# From Rapid Gains to Stalling: Two Decades of Modern Contraceptive Prevalence Rate in Ethiopia

**DOI:** 10.1101/2025.11.23.25340840

**Authors:** Yared Mekonnen, Daniel S Telake

**Author notes:** Corresponding Author: Email addresses: YM.

## Abstract

**Background:** Ethiopia, Africa’s second-most populous country, still faces high fertility and elevated maternal and child mortality. Although the country made substantial progress in expanding family planning uptake over the past decade, recent changes in contraceptive use trends remain insufficiently explored. This study uses nationally representative data to examine long-term trends in the modern contraceptive prevalence rate (mCPR).

**Method:** We analyzed mCPR trends among married women aged 15–49 using data from 10 national surveys (Demographic and Health Surveys [DHS] and Performance Monitoring for Action [PMA] surveys) conducted between 2000 and 2023. Piecewise regression models, adjusted for socio-demographic changes, were used to compare mCPR growth rates before and after 2015—the breakpoint identified by data visualization and confirmed by Chow test.

**Results:** Ethiopia’s mCPR rose from 6.2% in 2000 to 36.2% in 2015, demonstrating significant progress during that time. However, after 2015, the growth rate slowed significantly, with mCPR recorded at 36.6% in 2023. The annual growth rate during post-2015 period was 0.37 percentage points, compared to 2.1 percentage points annually pre-2015 (p=0.000). In rural areas, mCPR rose from 3.2% in 2000 to 32.7% in 2015 and 33.5% in 2023. The corresponding rates in urban areas were 29% in 2000, 49.6% in 2015, and 44.6% in 2023. In rural areas, annual growth dropped from 2.2 percentage points pre-2015 to 0.39 percentage points post-2015 (p=0.000), while urban areas saw a reversal trend, with growth falling from 1.82 percentage points annually pre-2015 to a negative rate of -0.56 percentage points post-2015 (p=0.000). The multivariate analysis reveals mCPR stall remains evident even after adjusting for socio-demographic factors.

**Conclusion:** Following a period of steady growth, Ethiopia’s progress in increasing mCPR has recently slowed significantly, with national rates stalling below 40% and declining in urban areas. This stall underscores the pressing need for renewed family planning efforts to address unmet need and overcome persistent barriers, including sociocultural challenges, equitable access limitations, funding issues, and service delivery gaps, in order to revive mCPR momentum and sustain progress.

## Background

Expanding access to contraception and meeting the demand for family planning through effective contraceptive methods are crucial steps toward achieving universal access to reproductive healthcare, as outlined in the 2030 Agenda for Sustainable Development [1]. Family planning is vital for reproductive health, providing numerous benefits for women, children, and families. Modern contraceptives help prevent unintended pregnancies, reduce unsafe abortions, and lower pregnancy-related health risks [2, 3]. By enabling proper birth spacing, family planning enhances child survival rates and gives mothers time to recover [4, 5]. It also empowers women by opening up opportunities for education and employment, thereby promoting gender equality and socioeconomic development [6]. Furthermore, lower fertility rates alleviate pressure on healthcare systems and social services, benefiting both communities and nations [7,8].

Globally, the use of modern contraceptives has steadily increased over the past few decades, contributing significantly to improved reproductive health outcomes. According to the United Nations, the global contraceptive prevalence rate (CPR) among married women of reproductive age rose from approximately 54% in 1990 to nearly 63% in 2020 [9]. However, regional disparities remain substantial. In high-income regions such as North America and Europe, modern contraceptive use exceeds 65% [2, 9]. In contrast, low- and middle-income countries, especially those in parts of Asia and Latin America, continue to face challenges in achieving higher mCPR levels, with rates ranging from 50% to 60% [10]. In sub-Saharan Africa, the mCPR has seen notable improvements but still falls short of global averages. From 2000 to 2020, the mCPR in the region increased from 13% to approximately 29% [11]. Despite this progress, sub-Saharan Africa remains the region with the lowest mCPR, primarily due to factors such as limited access to healthcare services, socio-cultural barriers, and a lack of awareness about modern contraceptive methods [4]. Within the region, there is significant variation; countries like South Africa and Kenya have achieved mCPR rates of over 60%, while others, such as Chad and the Democratic Republic of Congo, continue to have rates below 20% [11, 12]. This increase in mCPR in certain countries can be attributed to efforts like community-based distribution of contraceptives, the expansion of family planning services, and education initiatives. Nevertheless, the unmet need for contraception remains high in many sub-Saharan African countries, with approximately 24% of married or in-union women of reproductive age having an unmet need for modern contraception in 2020 [9, 10].

Ethiopia has made significant progress in its family planning programs over the past two decades, resulting in considerable increases in contraceptive use across the country. The Ethiopian government, in partnership with various stakeholders, has implemented a series of interventions aimed at enhancing access to and utilization of modern contraceptive methods. Through the Health Extension Program (HEP), which was launched in 2003, thousands of health extension workers have been trained and deployed to provide family planning services and other services, directly at the community level, including counseling, distribution of contraceptives, and referrals for long-term and permanent methods [13]. This community-based approach has been particularly effective in increasing awareness and uptake of family planning in rural areas, where access to health facilities has traditionally been limited. In addition to the HEP, the government has developed and implemented several policies to strengthen family planning services. As a result, Ethiopia’s mCPR increased from a mere 6.2% in 2000 to over 35% by 2016 [14]. This substantial growth in mCPR has contributed to a decline in total fertility rates (TFR) from 5.5 children per woman in 2000 to 4.6 in 2016, along with reductions in maternal and child mortality rates [14]. However, despite these achievements, challenges persist, such as disparities in contraceptive use across regions and population groups, particularly among rural and less-educated populations. Additionally, issues like poor method mix, high unmet need for family planning, commodity stockouts, and logistical difficulties continue to hinder progress [15, 16].

Studies on mCPR trends in Ethiopia have primarily focused on short-term trends, often overlooking long-term and recent trends, partly due to the lack of up-to-date data. Most of these studies, including those by Worku et al. (2015), Bogale et al. (2019), and Meselu et al. (2022), examine changes over five- or ten-year intervals using data from Demographic and Health Surveys [17–19], providing snapshots of contraceptive use during specific periods. Saifuddin et al. (2019) focused on more recent trends, identifying areas of progress and stagnation [20]. While these studies offer valuable insights, they do not present a comprehensive understanding of long-term mCPR trends in Ethiopia. Analyzing long-term trends is crucial to grasp the trajectory and pace of change in contraceptive use over time. Additionally, understanding these trends is essential for policymakers to evaluate Ethiopia’s progress toward meeting national and international family planning targets [21].

This study aims to examine mCPR trends in Ethiopia over a period of more than 20 years, from 2000 to 2023. This analysis utilizes data from two key national surveys: the Ethiopia Demographic and Health Surveys (EDHS) and the Performance Monitoring for Action (PMA) surveys. The availability of extensive data from both EDHS and PMA provides a unique opportunity to examine long-term trends not only at the national level but also by disaggregating the data into rural and urban areas and across various socio-demographic characteristics.

## Methods

### Data Sources and Inclusion Criteria

This study is based on an analysis of national data from 10 rounds of surveys (from 2000 to 2023) sourced from the Ethiopia Demographic and Health Surveys (EDHS) and the Ethiopia Performance Monitoring and Action (EPMA) surveys. The surveys were selected after a comprehensive review of all available national and sub-national household surveys in Ethiopia, in consultation with the Central Statistical Agency (CSA) and other organizations responsible for managing national data.

The criteria for including surveys in this analysis were: national or sub-national representativeness, similar sampling design, the use of a common national sampling frame for cluster selection, sample selection by the CSA, comparability of survey questionnaires for key variables, and availability of raw data. Surveys were not excluded based on their date of implementation as long as they met these criteria. Conversely, any surveys that did not fulfill these criteria were excluded from the analysis.

The EDHS is part of the worldwide MEASURE DHS project, funded by the United States Agency for International Development (USAID) and other development partners, with technical assistance provided by ICF International [14]. The first EDHS was conducted in 2000, and since then, the surveys were intended to be conducted at five-year intervals. So far, four EDHS were conducted in Ethiopia. However, the most recent EDHS data available is from 2016, indicating that the regular five-year interval was not maintained. In this analysis EDHS 2000, 2005, 2011 and 2016 were included.

The EPMA surveys are conducted annually to provide up-to-date and reliable information on population dynamics, reproductive health, family planning, and maternal health, addressing the lengthy gaps between successive DHS rounds [22]. The first EPMA survey was conducted in 2014, followed by successive annual surveys until 2023. The EPMA is led by the School of Public Health at Addis Ababa University in collaboration with regional universities, the Federal Ministry of Health, and the CSA. The Bill & Melinda Gates Institute for Population and Reproductive Health at Johns Hopkins Bloomberg School of Public Health provides overall direction and support, funded by the Bill & Melinda Gates Foundation. For this analysis, EPMA surveys from 2014, 2015, 2017, 2018, 2019, and 2023 met the inclusion criteria and were included.

Both the EDHS and EPMA surveys use nationally representative multi-stage cluster sampling, with the country’s census enumeration areas serving as clusters and primary sampling units. These surveys are designed to provide estimates for a range of indicators at the national level and separately for urban and rural areas and regions. The CSA was responsible for selecting sample clusters using the standard DHS sampling methodology, ensuring comparability between the two surveys. The raw data, along with detailed data dictionaries and data structures, are publicly available upon request. By pooling data from these surveys, we obtained a large dataset of trends to allow for robust statistical analyses with high precision.

### Study Variables

This study focuses on several key variables, including the modern contraceptive prevalence rate (mCPR), survey year, residence (urban-rural), marital status, women’s age, number of children ever born, education level, household wealth, and other socio-economic characteristics. These variables were gathered in a similar manner in both the EDHS and EPMA surveys. The primary outcome variable is mCPR, defined as the proportion of married women aged 15–49 who are currently using, or whose partners are using, a modern method of contraception at the time of the survey.

### Data pooling and categorization

The data from the 10 surveys were pooled to create a large serial cross-sectional dataset for this analysis. Women’s age was categorized into four groups: 15–19, 20–24, 25–34, and 35–49 years. The number of children ever born was categorized into six groups: no child, 1, 2, 3, 4, and 5 or more. Education status was divided into three categories: no education, elementary education (1–6 years of schooling), and secondary or higher education (7 or more years of schooling). The EDHS and EPMA survey datasets provided a wealth index constructed using principal component analysis, which ranked households into five wealth quintiles: first, second, middle, fourth, and highest [23].

### Data analysis

This study used piecewise regression to examine mCPR trends, estimating slopes before and after a specified breakpoint year (the “knot”). The analysis was conducted at both the national level and separately for urban and rural areas to explore variations by residence. The piecewise regression model was specified as follows:

**mCPR = β_0_ + β_1_(year_pre_knot) + β_2_(year_post_knot) + β_3_Z + ɛ**

Where:

– **mCPR**: modern Contraceptive Prevalence Rate (dependent variable).
– **year_pre_knot**: a continuous variable representing the general time trend before the “knot” (breaking year).
– **year_post_knot**: a continuous variable representing the years after the “knot” (set to 0 for years before the “knot”).
– **Z**: a vector of covariates (including age, education, number of children and wealth).
– **β_0_**: Intercept.
– **β_1_**: coefficient for the time trend before the “knot”.
– **β₂:** coefficient for the change in trend after the “knot”.
– **β_1_ - β₂** represents the change in the growth rate of mCPR after the “knot” compared to the period before the “knot”. If **β₂** is greater than **β_1_**, it indicates an acceleration in mCPR growth post the “knot”. If **β₂** is smaller, it suggests a slowdown or plateau in the growth rate after the “knot”.
– **β_3_**: a vector of coefficients for the covariates.
– **ɛ:** error term.

We tested for structural breaks in mCPR trends using Chow tests within a survey-weighted linear regression. The Chow test assesses whether regression coefficients differ across two subsets split at a pre-specified breakpoint year (the “knot”) by comparing a single linear model (no break) to two segments-specific models (pre- and post-knot). Under the null (no break), both the level and slope change terms are zero (β2=0, β3=0); the alternative allows β2≠0 and/or β3≠0, indicating a shift in level and/or trend. The test statistic follows an F-distribution with 2 and (n−4) degrees of freedom. We evaluated candidate break years and selected the “knot” with the largest F-statistic, providing the strongest evidence of a break while limiting post-hoc selection bias.

We used Stata version 18 (Stata Corporation, College Station, TX, USA) for data management and analyses. The Survey command in STATA was used to delineate the strata and primary sampling unit.

## Result

### Compositional changes in socio-demographic characteristics of the women

Table 1 presents the distribution of key socio-demographic characteristics among married women aged 15–49 years over the period 2000–2023. This analysis examines temporal patterns in the socio-demographic profile of these women, focusing on age, parity, education, and household wealth across the 23-year span. These characteristics are known to influence contraceptive use in Ethiopia and other sub-Saharan African settings, and any compositional shifts in the population—such as changes in the proportion of women with higher education or varying parity—can affect modern contraceptive prevalence rate (mCPR) trends independently of behavioral factors that directly drive contraceptive uptake. It is therefore imperative to understand the trends in these characteristics and account for observed changes when evaluating and estimating mCPR trends.

**Table 1.** Selected background characteristics of married women age 15-19 years, Ethiopia, 2000–2023.

| Characteristic | 2000<br>(EDHS)<br>N=9,297 | 2005<br>(EDHS)<br>N=8,438 | 2011<br>(EDHS)<br>N=9,478 | 2014<br>(EPMA)<br>N=3,672 | 2015<br>(EPMA)<br>N=4,240 | 2016<br>(EDHS)<br>N=9,602 | 2017<br>(EPMA)<br>N=4,292 | 2018<br>(EPMA)<br>N=4,217 | 2019<br>(EPMA)<br>N=5,488 | 2023<br>(EPMA)<br>N=5,565 |
| --- | --- | --- | --- | --- | --- | --- | --- | --- | --- | --- |
| **Residence** |  |  |  |  |  |  |  |  |  |  |
| Urban | 18.2 | 17.8 | 23.9 | 23.1 | 23.1 | 22.2 | 24.6 | 25.3 | 25.8 | 29.7 |
| Rural | 81.8 | 82.2 | 76.1 | 76.9 | 76.9 | 77.8 | 75.5 | 74.7 | 74.2 | 70.3 |
| **Age Group** |  |  |  |  |  |  |  |  |  |  |
| 15–19 | 24.1 | 23.2 | 24.3 | 23.5 | 23.5 | 21.6 | 23.7 | 23.4 | 22.4 | 21.9 |
| 20–24 | 18.6 | 18.1 | 17.8 | 18.3 | 18.7 | 17.6 | 16.5 | 16.6 | 17.5 | 17.4 |
| 25–34 | 28.7 | 30.7 | 31.5 | 33.0 | 31.2 | 33.8 | 33.6 | 32.4 | 31.8 | 31.8 |
| 35–49 | 28.5 | 28.0 | 26.5 | 25.2 | 26.5 | 27.0 | 26.2 | 27.6 | 28.4 | 28.9 |
| **Children Ever Born** |  |  |  |  |  |  |  |  |  |  |
| 0 | 31.9 | 31.0 | 33.8 | 33.7 | 33.7 | 32.5 | 32.8 | 34.8 | 32.6 | 34.8 |
| 1 | 10.6 | 10.4 | 11.0 | 11.9 | 12.9 | 11.7 | 12.5 | 12.1 | 14.1 | 13.2 |
| 2 | 10.4 | 9.6 | 10.7 | 11.1 | 10.2 | 10.2 | 10.3 | 11.0 | 12.1 | 13.2 |
| 3 | 8.7 | 9.0 | 8.7 | 9.3 | 9.2 | 9.6 | 9.5 | 9.0 | 8.6 | 10.4 |
| 4 | 7.2 | 8.5 | 8.2 | 8.9 | 8.5 | 8.2 | 9.1 | 8.6 | 7.9 | 8.2 |
| 5+ | 31.3 | 31.6 | 27.8 | 25.1 | 25.7 | 27.9 | 25.8 | 24.6 | 24.8 | 20.2 |
| **Education** |  |  |  |  |  |  |  |  |  |  |
| No education | 75.3 | 65.9 | 50.8 | 44.3 | 44.5 | 47.8 | 41.6 | 38.2 | 37.7 | 28.4 |
| Primary | 15.6 | 22.2 | 38.0 | 37.2 | 37.1 | 35.0 | 37.9 | 39.7 | 36.7 | 40.7 |
| Secondary+ | 9.0 | 11.9 | 11.2 | 18.6 | 18.4 | 17.2 | 20.5 | 22.1 | 25.6 | 30.9 |
| **Wealth Quintile** |  |  |  |  |  |  |  |  |  |  |
| Lowest | 11.8 | 17.3 | 18.1 | 17.3 | 20.4 | 16.8 | 20.2 | 20.2 | 19.3 | 18.6 |
| Second | 14.7 | 18.8 | 18.4 | 19.1 | 20.0 | 17.9 | 19.2 | 19.1 | 19.3 | 19.0 |
| Middle | 20.6 | 19.4 | 18.4 | 20.4 | 19.4 | 19.0 | 18.6 | 18.8 | 19.3 | 19.2 |
| Fourth | 19.8 | 18.8 | 19.5 | 20.3 | 18.6 | 19.8 | 19.9 | 19.2 | 19.4 | 19.7 |
| Highest | 33.1 | 25.7 | 25.7 | 22.9 | 21.7 | 26.6 | 22.0 | 22.7 | 22.7 | 23.5 |

Over the 23-year period, urban residence increased from 18.2% in 2000 to 29.7% in 2023, with a corresponding decline in rural residence from 81.8% to 70.3%, reflecting accelerated urbanization. Age distributions showed a modest decrease in the proportion of women aged 15–19 years (from 24.1% to 21.9%), stability in the 20–24 age group (around 17–18%), a slight increase in the 25–34 group (from 28.7% to 31.8%), and relative constancy in the 35–49 group (approximately 28%). Parity patterns indicated a rise in nulliparous women (from 31.9% to 34.8%) and low-parity groups (e.g., one child: from 10.6% to 13.2%; two children: from 10.4% to 13.2%), alongside a marked decline in high parity (five or more children: from 31.3% to 20.2%), with the reduction accelerating post-2011. Educational attainment improved substantially, with the proportion of women with no education dropping from 75.3% to 28.4%, primary education rising from 15.6% to 40.7%, and secondary or higher education increasing from 9.0% to 30.9%, showing consistent gains especially after 2005. Household wealth distributions shifted toward greater equity, as the highest quintile decreased from 33.1% to 23.5%, while lower quintiles (lowest and second) saw modest increases (e.g., lowest: from 11.8% to 18.6%). Overall, these compositional changes highlight evolving demographic profiles that may influence mCPR trends independently of behavioral factors like attitudes or service access. Thus, the piecewise regressions presented here incorporate these covariates to isolate compositional effects from true changes, enhancing trend accuracy.

### Observed mCPR trends, and structural break in mCPR trends

From 2000 to 2023, Ethiopia’s modern contraceptive prevalence rate (mCPR) rose nearly six-fold, from 6.2% to 36.6%. Rapid gains occurred before 2011, followed by marked deceleration and stagnation around 35–37% (Table 2). Urban mCPR climbed from 29.0% in 2000 to a peak of 49.6% in 2015, then plateaued and declined slightly to 44.6% by 2023, indicating clear stagnation and possible reversal. In contrast, rural mCPR increased more than ten-fold, from 3.2% to 33.5%, with the fastest rise before 2014 and stabilization or slow growth thereafter. Consequently, the urban–rural gap narrowed from 25.8 to 11.1 percentage points; recent national stagnation primarily reflects arrested urban progress, while rural areas show stabilization or slow growth.

**Table 2.** Trends (observed) in mCPR (%) for the total, by urban-rural and selected socioeconomic characteristics, 2000-2023, Ethiopia.

| Data source | 2000<br>(EDHS)<br>N=9,297 | 2005<br>(EDHS)<br>N=8,438 | 2011<br>(EDHS)<br>N=9,478 | 2014<br>(EPMA)<br>N=3,672 | 2015<br>(EPMA)<br>N=4,240 | 2016<br>(EDHS)<br>N=9,602 | 2017<br>(EPMA)<br>N=4,292 | 2018<br>(EPMA)<br>N=4,217 | 2019<br>(EPMA)<br>N=5,488 | 2023<br>(EPMA)<br>N=5,565 |
| --- | --- | --- | --- | --- | --- | --- | --- | --- | --- | --- |
| **mCPR** |  |  |  |  |  |  |  |  |  |  |
| Ethiopia | 6.2 | 13.7 | 26.5 | 34.0 | 35.7 | 35.2 | 34.8 | 37.6 | 34.8 | 36.6 |
| **Residence** |  |  |  |  |  |  |  |  |  |  |
| Urban | 29.0 | 41.1 | 49.7 | 46.6 | 49.6 | 49.5 | 44.3 | 44.0 | 43.9 | 44.6 |
| Rural | 3.2 | 10.6 | 21.7 | 31.4 | 32.7 | 32.5 | 32.7 | 36.0 | 31.6 | 33.5 |

The marked flattening of the national trend from the mid-2010s onward prompted formal testing for a structural break (knot). Using Chow tests, we evaluated potential breakpoints (“knot”) at 2015, 2016, and 2017 (Table 3). All tested knots revealed statistically significant structural breaks nationally (F = 6.7–7.9, p =0.002) and in both urban and rural strata (p < 0.005). Pre-break unadjusted annual increases of approximately 2 percentage points slowed dramatically post-break, most evidently with the 2015 knot (national slope from +1.99 to +0.14 pp/year). Urban areas showed the strongest evidence of a break (F ≥ 10.0, p < 0.001), shifting from modest gains to stagnation or decline (−0.63 pp/year after 2015). Rural deceleration was less severe but still significant, with growth slowing from approximately 2 pp/year to near zero. The 2015 knot consistently yielded the highest F-statistics across strata and the sharpest contrast in slopes, particularly the reversal in urban areas, and was therefore selected as the optimal breakpoint for subsequent segmented analyses.

**Table 3.** Testing structural breaks (knots) in mCPR trends for Ethiopia and by residence.

|  | **Knot**<br>**(Year)** | **F_stat** | **Unadjusted**<br>**Pre-knot slope**<br>**(pp changes/year)** | **Unadjusted**<br>**Post-knot slope**<br>**(pp changes /year)** | **P-value** |
| --- | --- | --- | --- | --- | --- |
| **Ethiopia** | 2015 | 7.9 | 1.99 | 0.14 | 0.001 |
|  | 2016 | 7.0 | 2.02 | 0.18 | 0.002 |
|  | 2017 | 6.7 | 1.95 | 0.16 | 0.002 |
| **Urban** | 2015 | 10.4 | 1.34 | -0.63 | 0.000 |
|  | 2016 | 10.0 | 1.26 | -0.42 | 0.000 |
|  | 2017 | 10.1 | 1.19 | 0.08 | 0.000 |
| **Rural** | 2015 | 7.2 | 1.96 | 0.08 | 0.002 |
|  | 2016 | 5.4 | 2.01 | 0.06 | 0.005 |
|  | 2017 | 5.4 | 1.97 | -0.04 | 0.005 |

*Notes: pp= percentage points. F-statistics are based on the Chow test for joint changes in intercept and slope. p-Values indicate significance of the break; lower values suggest stronger evidence against the null hypothesis of no break.*

### Piecewise regression of mCPR growth rate: pre-post 2015 changes

Table 4 presents results from piecewise regression models—adjusted for women’s age, parity, education, and household wealth—showing the annual rate of change in modern contraceptive prevalence rate (mCPR) before and after 2015. These models examine the change in the rate of increase (or slope) of mCPR trends for Ethiopia overall, as well as for rural and urban areas.

**Table 4:** Piecewise Regression adjusted coefficients of mCPR Changes and 95% Confidence Interval (CI) Pre- and Post-2015, models adjusted for socio-demographics, National, Urban, and Rural Ethiopia.

|  | Ethiopia |  |  |  | Rural |  |  |  | Urban |  |  |  |
| --- | --- | --- | --- | --- | --- | --- | --- | --- | --- | --- | --- | --- |
| | β-Coef. | p-value | 95% CI | | β-Coef. | p-value | 95% CI | | β-Coef. | p-value | 95% CI | |
|  |  |  | Lower | Upper |  |  | Lower | Upper |  |  | Lower | Upper |
| **Year** |  |  |  |  |  |  |  |  |  |  |  |  |
| Pre-2015 | 0.019 | 0.000 | 0.018 | 0.022 | 0.021 | 0.000 | 0.019 | 0.023 | 0.016 | 0.000 | 0.011 | 0.021 |
| Post-2015 | -0.020 | 0.000 | -0.028 | -0.013 | -0.019 | 0.000 | -0.028 | -0.010 | -0.021 | 0.000 | -0.030 | -0.012 |
| **Age** |  |  |  |  |  |  |  |  |  |  |  |  |
| 15-19 | 0.000 |  |  |  | 0.000 |  |  |  | 0.000 |  |  |  |
| 20-24 | 0.006 | 0.542 | -0.014 | 0.027 | 0.013 | 0.248 | -0.009 | 0.034 | 0.028 | 0.387 | -0.035 | 0.091 |
| 25-34 | 0.007 | 0.575 | -0.018 | 0.033 | 0.035 | 0.008 | 0.009 | 0.061 | -0.058 | 0.063 | -0.118 | 0.003 |
| 35-49 | -0.028 | 0.074 | -0.060 | 0.003 | 0.025 | 0.096 | -0.005 | 0.055 | -0.231 | 0.000 | -0.302 | -0.160 |
| **Children ever born** |  |  |  |  |  |  |  |  |  |  |  |  |
| 0 | 0.000 |  |  |  | 0.000 |  |  |  | 0.000 |  |  |  |
| 1 | 0.144 | 0.000 | 0.121 | 0.166 | 0.107 | 0.000 | 0.081 | 0.133 | 0.249 | 0.000 | 0.213 | 0.286 |
| 2 | 0.164 | 0.000 | 0.139 | 0.188 | 0.108 | 0.000 | 0.080 | 0.136 | 0.337 | 0.000 | 0.300 | 0.374 |
| 3 | 0.150 | 0.000 | 0.124 | 0.177 | 0.096 | 0.000 | 0.068 | 0.124 | 0.336 | 0.000 | 0.291 | 0.382 |
| 4 | 0.140 | 0.000 | 0.114 | 0.167 | 0.083 | 0.000 | 0.056 | 0.111 | 0.333 | 0.000 | 0.278 | 0.387 |
| 5+ | 0.110 | 0.000 | 0.082 | 0.138 | 0.047 | 0.001 | 0.018 | 0.075 | 0.287 | 0.000 | 0.234 | 0.340 |
| **Education** |  |  |  |  |  |  |  |  |  |  |  |  |
| No education | 0.000 |  |  |  | 0.000 |  |  |  | 0.000 |  |  |  |
| Primary | 0.075 | 0.000 | 0.059 | 0.091 | 0.072 | 0.000 | 0.055 | 0.090 | 0.055 | 0.001 | 0.022 | 0.089 |
| Secondary + | 0.135 | 0.000 | 0.111 | 0.158 | 0.167 | 0.000 | 0.133 | 0.202 | 0.097 | 0.000 | 0.062 | 0.133 |
| **Household Wealth** |  |  |  |  |  |  |  |  |  |  |  |  |
| lowest | 0.000 |  |  |  | 0.000 |  |  |  | 0.000 |  |  |  |
| Second | 0.043 | 0.000 | 0.025 | 0.061 | 0.041 | 0.000 | 0.023 | 0.060 | 0.076 | 0.178 | -0.035 | 0.186 |
| Middle | 0.076 | 0.000 | 0.058 | 0.095 | 0.077 | 0.000 | 0.057 | 0.096 | 0.049 | 0.271 | -0.038 | 0.136 |
| Fourth | 0.103 | 0.000 | 0.082 | 0.124 | 0.110 | 0.000 | 0.088 | 0.131 | 0.029 | 0.523 | -0.061 | 0.120 |
| Highest | 0.118 | 0.000 | 0.093 | 0.143 | 0.129 | 0.000 | 0.103 | 0.155 | 0.083 | 0.090 | -0.013 | 0.180 |
| **Residence** |  |  |  |  |  |  |  |  |  |  |  |  |
| Urban | 0.000 |  |  |  |  |  |  |  |  |  |  |  |
| Rural | -0.074 | 0.000 | -0.104 | -0.045 |  |  |  |  |  |  |  |  |
| *Models are piecewise regression that divide the trend into two segments: one covering the period before 2015 and after 2015* |  |  |  |  |  |  |  |  |  |  |  |  |

For Ethiopia as a whole, the pre-2015 period exhibited a statistically significant positive trend in mCPR growth. The unadjusted model indicates an annual increase of 2.06 percentage points (β = 0.0206, p < 0.001), while the adjusted model shows a similar positive trend with an annual increase of 1.98 percentage points (β = 0.0198, p < 0.001). However, after 2015, there is a marked and significant slowdown in mCPR growth. The coefficient for this period turns negative in both the unadjusted (β = -0.0169, p < 0.000) and adjusted models (β = -0.0204, p < 0.001).

Comparing the coefficients between the pre-2015 and post-2015 periods reveal the actual rate of mCPR growth after 2015. Specifically, the annual increase in mCPR dropped significantly to just 0.37 percentage points in the unadjusted model (2.06% minus 1.69%) and turned negative at - 0.06 percentage points in the adjusted model (1.98% minus 2.04%). These findings indicate a substantial deceleration and stalling in mCPR growth starting from 2015.

In rural Ethiopia, there are notable changes in mCPR growth trends before and after 2015. Before 2015, the unadjusted model shows a significant annual increase in mCPR of 2.2 percentage points (β = 0.0216, p < 0.001), which is consistent with the adjusted model (β = 0.0209, p < 0.001). However, psot-2015, there is a significant slowdown in the growth rate. The rate of change for this period turns significantly negative in both the unadjusted (β = -0.0177, p = 0.002) and adjusted (β = -0.0186, p < 0.001) models, indicating a marked change compared to the pre-2015 trend. In the unadjusted rural model, the annual mCPR increase drops to 0.39 percentage points per year (i.e., 2.16% minus 1.77%) after 2015. In the adjusted rural model, the rate decreases to just 0.23 percentage points per year (i.e., 2.09% minus 1.86%), suggesting an almost stable growth rate in mCPR during this period.

In urban Ethiopia, the pre-2015 mCPR growth rate was slower compared to rural areas, and a more pronounced decline occurred after 2015. Before 2015, the unadjusted model shows an annual mCPR increase of 1.82 percentage points (β = 0.0182, p < 0.001), while the adjusted model indicates a slightly lower annual increase of 1.57% (β = 0.0157, p < 0.001). However, after 2015, there is a sharp reversal in the growth rate. The slope turns significantly negative in both models: unadjusted model (β = -0.0217, p < 0.001) and adjusted model (β = -0.0192, p < 0.001). These findings indicate a negative mCPR growth rate post-2015 in urban Ethiopia, resulting in a -0.56-percentage points deceleration in the unadjusted model and a -0.53-percentage points deceleration in the adjusted model. This highlights a significant reversal in mCPR growth in urban areas starting from 2015.

In addition, the multivariate models reveal that mCPR is positively associated with higher parity (strongest for 1–4 children, β = 0.140–0.164 nationally; more pronounced in urban areas), higher educational attainment (β = 0.135 for secondary+, strongest in rural areas), and greater household wealth (positive gradient, clearest in rural settings). In contrast, women aged 35–49 years and rural residence show lower mCPR (β = -0.028 nationally, strongly negative in urban areas for older women; β = -0.074 for rural residence). Socioeconomic gradients are generally stronger in rural than urban areas, with urban effects often weakened or non-significant for education and wealth.

### Predicted mCPR trends: 2000-2025

The predicted values of mCPR at the national, rural, and urban levels for the period 2000–2025 are illustrated in Figure 1a-c. These predictions are derived from the adjusted piecewise regression models, which segment the trends into two distinct periods: pre-2015 and post-2015. This approach allows the model to capture changes in the growth rate of mCPR over time. The predicted values represent the model’s estimation of how mCPR changes year-by-year, based on historical data and observed shifts in trends. Prior to 2015, the model clearly captures a period of steady growth in mCPR. However, after 2015, the predicted values reveal a noticeable stall in mCPR, suggesting a slowdown in the progress of contraceptive use. The model closely mirrors the observed rise in mCPR up to 2015 and accurately depicts the subsequent plateau in the years that follow.

**Figure 1a-c.**
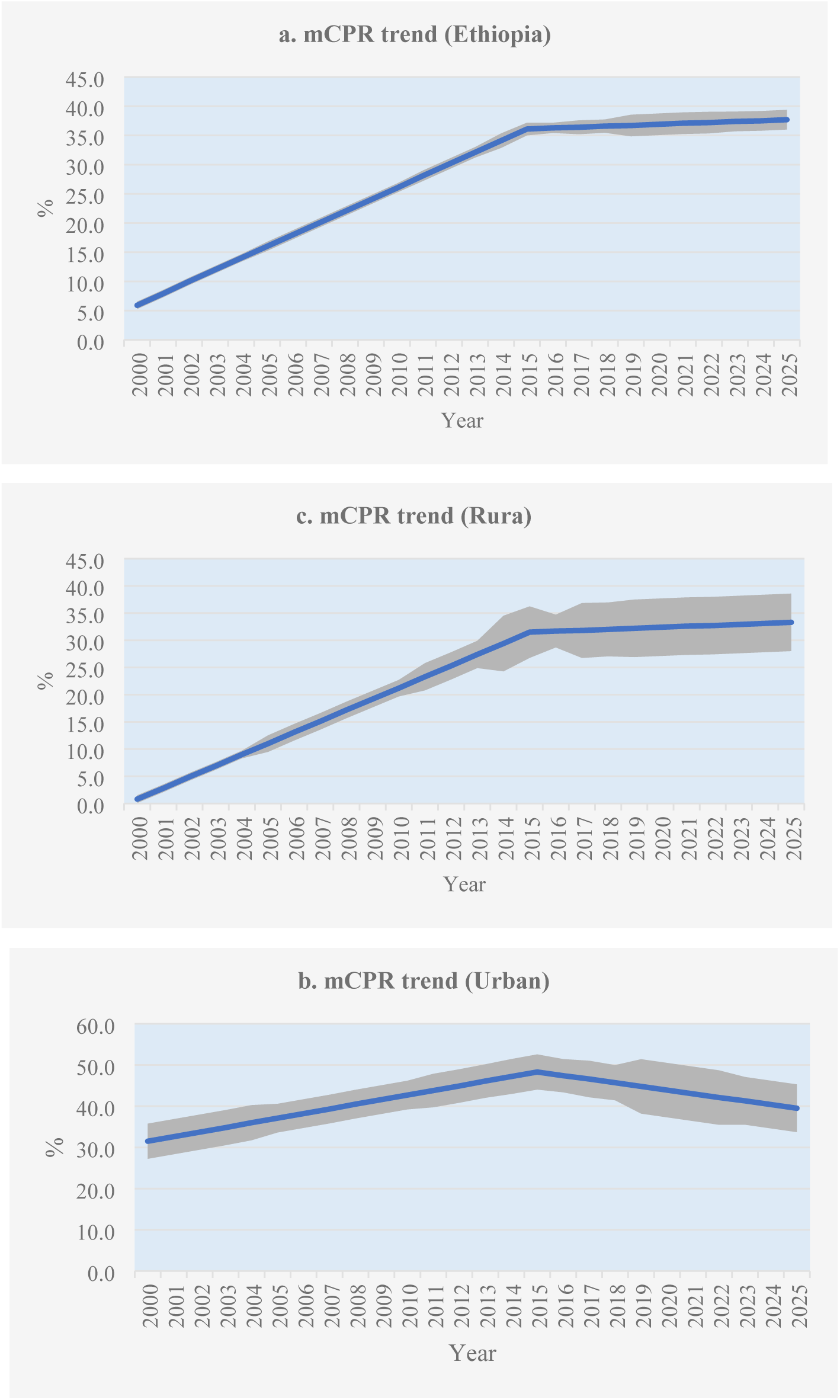
Predicted mCPR (%) trends (using piecewise regression), 2000-2025: (a) Ethiopia, (b) rural, and (c) urban.

## Discussion

The findings of this study reveal a significant stall in the growth of mCPR in Ethiopia in recent years, following a period of steady progress from the mid-2000s to the mid-2010s. After 2015, our analysis showed that the mCPR growth rate slowed to 0.37 percentage points nationally and 0.39 percentage points in rural areas, with a notable decline of -0.56 percentage points in urban areas. Multivariate analysis further confirms that these trends persisted even after adjusting for socio-demographic factors. According to the literature, a stall in mCPR is defined as a period when the annual growth rate in contraceptive use significantly slows, falling below a threshold usually set between 0 and 0.6 percentage points per year [4, 24]. In line with this definition, Ethiopia’s current mCPR trends can indeed be characterized as experiencing a stall, with urban areas even showing a declining trend. Understanding these stalls is crucial to identifying the barriers impeding progress and developing strategies to overcome them, thereby reigniting efforts to improve contraceptive use in Ethiopia.

Stalls in mCPR are not unique to Ethiopia; similar trends have been observed across various African and developing countries. Earlier multi-country studies have explored the occurrence and characteristics of these stalls. Ross et al. (2004) analyzed contraceptive prevalence plateaus across 52 countries using data from national surveys [25]. They identified plateaus in countries at different stages of contraceptive adoption, across regions such as sub-Saharan Africa, South Asia, and Latin America. Many stalls occurred at low levels of contraceptive use, indicating challenges in establishing a steady upward trajectory. Several sub-Saharan African countries experienced mCPR plateaus between 2000 and 2019, most of which occurred at levels below 20% [24]. These varying stall levels indicate notable stagnation or slowdown in contraceptive growth, highlighting the complex challenges in maintaining a consistent upward trajectory in mCPR. In some instances, stalls were observed at intermediate levels of prevalence (25-60%), suggesting a slowdown even after a period of growth. In countries with higher initial mCPR, mCPR stall often signaled a saturation point. For instance, Kenya exhibited a plateauing in contraceptive use in recent years, with mCPR reached a notable high of 62.3% in 2015 and stall at around 60% from 2018 to 2020. Although this rate is higher than the less than 40% level found in our analysis, it still indicates a stall at a much higher level than previously observed [26]. Similarly, South Africa experienced a stall from 2009 to 2013, hovering around the 60% threshold [24].

Ethiopia’s stall in mCPR occurred at a level of less than 40%, which is relatively lower than the plateau levels documented in countries like Kenya and South Africa, where mCPR reached around 60%. However, Ethiopia’s stall occurred at a higher rate compared to several West African countries, which experienced plateaus at mCPR levels below 20% [24]. Typically, contraceptive stalls are more commonly observed when countries reach higher mCPR levels, often above 50%. At these higher levels, further increases in contraceptive use often become more challenging due to a combination of factors such as reduced unmet need, social and cultural barriers, and the difficulties in reaching marginalized and hard-to-reach populations.

The reasons for the contraceptive stall in Ethiopia are not fully understood; however, insights can be drawn from similar trends observed in other sub-Saharan African countries. Studies in the region points to a complex mix of factors affecting both the demand and supply sides of family planning services, which may also apply to Ethiopia’s situation. On the demand side, one key factor is resistance to modern contraception, often rooted in traditional beliefs, religious views, and cultural norms. In many communities, these deep-seated attitudes foster misconceptions about contraceptive methods, including concerns about potential side effects or fears that modern contraception could affect fertility [27]. Such cultural resistance can deter individuals, particularly women, from seeking family planning services, even when they express a desire to limit or space pregnancies. On the supply side, several programmatic shortcomings can hinder the effective delivery of family planning services. A primary issue is the lack of resources dedicated to family planning programs. Insufficient funding affects the availability of contraceptives and limits the reach of services, particularly in remote or underserved areas [24]. Additionally, supply chain disruptions, such as stockouts of contraceptive supplies, can prevent consistent access to the necessary products. Poor logistics, and inadequate distribution systems exacerbate these supply chain issues, leading to gaps in service delivery [20].

As in other sub-Saharan African countries, Ethiopia’s family planning program faces persistent challenges that may have contributed to the recent stall in the mCPR. Long-standing equity gaps due to geographic and sociodemographic disparities limit access to family planning services [28]. Supply chain management issues add further complications, particularly in remote and hard-to-reach areas. Despite increased domestic financing for family planning, the program still heavily relies on external funding, raising concerns about its long-term sustainability [29]. Additionally, social and cultural barriers, such as gender inequality and misconceptions about contraceptives, continue to impede progress [30]. The quality of family planning services is often inadequate, which affects the uptake and sustained use of contraceptives [31]. Revitalizing the momentum in Ethiopia’s mCPR requires addressing these challenges, as there is still potential for growth. With about 15% of married women in Ethiopia having an unmet need for family planning in 2023, a strong program can further increase contraceptive use [32].

Review of available literature suggest that addressing the contraceptive stall requires a multifaceted approach. Increasing investment in family planning is crucial for strengthening supply chains and preventing stockouts [24]. Governments should focus on improving access to a range of contraceptive methods, especially in underserved areas, and upgrading health facilities to ensure consistent service delivery. Engaging communities is vital for shifting norms and attitudes towards contraception; involving leaders, religious figures, and men in discussions can foster positive attitudes and support joint decision-making [27]. Effective communication through media, social marketing, and health workers can help raise awareness and promote family planning, particularly in regions experiencing stagnation [20, 27]. Expanding contraceptive options, such as implants and IUDs, and training health workers to offer diverse methods enable individuals to make informed choices [20]. Strong political commitment is also essential; governments must implement supportive policies, secure adequate funding, and conduct continuous monitoring to promptly address any stalls in progress [20, 24].

Finally, this study has several strengths that deserve mentioning, including the use of comprehensive, nationwide data from the Demographic and Health Surveys (DHS) and PMA data since 2000. By combining these sources, it establishes a strong foundation for analyzing contraceptive trends across Ethiopia’s rural and urban population, providing valuable population-level insights. However, the study has some limitations, including the absence of regional or subnational analysis due to small sample size issues, which could have revealed significant variations within Ethiopia’s regions. Furthermore, the trend analysis is based on 10 data points with uneven intervals, particularly before 2014; more frequent and evenly spaced data collection could have provided a clearer picture of annual mCPR growth and changes over time.

## Conclusion

This analysis of nationwide survey data reveals a significant stall in Ethiopia’s mCPR, affecting both rural and urban areas. Urban areas, in particular, show a noticeable decline. These trends align with patterns seen in other African countries. Ethiopia’s stall in mCPR, currently below 40%, suggests potential for growth, as a significant 15% of married women still have an unmet need for contraception. Tackling this stall and meeting the unmet need require a multifaceted approach, including increased investment in family planning, broader access to diverse contraceptive methods, community engagement to shift social norms, a focus on equitable access, effective communication strategies, and strong political commitment. Implementing these strategies can revive momentum in Ethiopia’s mCPR and ensure sustainable progress in family planning. Further research is warranted to identify the causes of the stall in both rural and urban settings to inform targeted interventions.

## List of abbreviations

CI: Confidence Interval
CSA: Central Statistical Agency
DHS: Demographic and Health Survey
EDHS: Ethiopia Demographic and Health Survey
EPMA: Ethiopia Performance Monitoring and Accountability
mCPR: Modern Contraceptive Prevalence Rate
NGO: Non-Governmental Organization
PMA: Performance Monitoring and Accountability
USAID: United States Agency for International Development.

## Declarations

### Ethical approval and consent to participate

Not applicable

### Consent to publish

Not applicable

### Data Availability

The EDHS data used in this study are openly accessible and can be downloaded at https://dhsprogram.com/data/available-datasets.cfm. The EPMA raw data were obtained upon permission and downloaded from [https://www.pmadata.org/data/about-data](https://www.pmadata.org/data/about-data). Both data accessed on April 17, 2025.

### Authors’ contributions

YM conceived and designed the study, analyzed the data and wrote the manuscript. DT were involved in the data analysis and interpretation of the data and wrote the manuscript. YM, DT approved the final manuscript.

## Acknowledgments

The DHS data were obtained from the ICF International, Calverton, Maryland, USA. We obtained the EPMA survey data upon request from PMA2020.

